# Effect of Long-Term Caloric Restriction on DNA Methylation Measures of Biological Aging in Healthy Adults: CALERIE™ Trial Analysis

**DOI:** 10.1101/2021.09.21.21263912

**Authors:** R Waziry, DL Corcoran, KM Huffman, MS Kobor, M Kothari, VB Kraus, WE Kraus, DTS Lin, CF Pieper, ME Ramaker, M Bhapkar, SK Das, L Ferrucci, WJ Hastings, M Kebbe, DC Parker, SB Racette, I Shalev, B Schilling, DW Belsky

**Affiliations:** Butler Columbia Aging Center, Columbia University Mailman School of Public Health, New York, NY, USA; Center for Genomic and Computational Biology, Duke University, NC, USA; Department of Medicine and Molecular Physiology Institute, Duke University School of Medicine, Durham, NC, USA; Department of Medical Genetics, BC Children’s Hospital Research Institute, Centre for Molecular Medicine and Therapeutics, University of British Columbia, Vancouver, BC, Canada; Department of Biostatistics and Bioinformatics, Duke University School of Medicine, Durham, NC, USA; US Department of Agriculture, Jean Mayer Human Nutrition Research Center on Aging at Tufts University, Boston, MA, USA; Translational Gerontology Branch, National Institute on Aging, National Institutes of Health, Baltimore, MD, USA; Department of Biobehavioral Health, Pennsylvania State University, University Park, PA, USA; Pennington Biomedical Research Center, Baton Rouge, LA, USA; Washington University School of Medicine, St. Louis, MO, USA; Buck Institute for Research on Aging, Novato, CA, USA; Department of Epidemiology, Columbia University Mailman School of Public Health, New York, NY, USA

**Keywords:** biological age, DNA methylation, caloric restriction, randomized clinical trial, geroprotector, geroscience

## Abstract

Calorie restriction (CR) slows aging and increases healthy lifespan in model organisms. We tested if CR slowed biological aging in humans using DNA methylation analysis of blood samples from N=197 participants in the Comprehensive Assessment of Long-term Effects of Reducing Intake of Energy (CALERIE™) randomized controlled trial. We quantified CR effects on biological aging by comparing change scores for six epigenetic-clock and Pace-of-Aging measures between n=128 CR-group and n=69 ad-libitum-control-group participants at 12- and 24-month follow-ups. CR effects were strongest for DunedinPACE Pace of Aging (12-month Cohen’s d=0.3; 24-month Cohen’s d=0.2, p<0.01 for both), followed by DunedinPoAm and the GrimAge epigenetic clock, although effects for these measures were not statistically different from zero (p>0.08). CR effects for other epigenetic clocks were in the opposite direction (all p>0.15). CALERIE intervention slowed Pace of Aging but showed minimal effect on epigenetic clocks hypothesized to reflect longer term accumulation of aging burden.

## INTRODUCTION

The geroscience hypothesis proposes that therapy to slow or reverse molecular changes that occur with aging can delay or prevent multiple chronic diseases and extend healthy lifespan ^1–3^. Animal experiments provide support for this hypothesis; behavioral interventions and drug therapies can improve cellular communication and nutrient sensing, regulate transcription, slow stem cell aging and telomere attrition, and extend healthy lifespan ^4–6^. Translation of such interventions to slow biological aging in humans is needed to address the increased burden of disease forecasted to accompany population aging ^7–9^.

Calorie restriction (CR), defined as lessening caloric intake without depriving essential nutrients ^10^, is established to increase healthy lifespan in multiple species, including non-human primates ^11–13^. The National Institute on Aging’s Comprehensive Assessment of Long-Term Effects of Reducing Intake of Energy (CALERIE™) Trial was conducted to test effects of long-term CR in healthy, non-obese humans ^14^. In CALERIE™, n=220 non-obese young-to-middle aged adults were randomized in a 2:1 ratio to either a prescribed a dose of 25% CR or ad libitum (AL) control diet for two years. Molecular analysis of CALERIE data have been equivocal, with no changes observed in some aging-related biomarkers ^14,15^. In contrast, analysis of clinical data from the CALERIE trial indicate intervention improved cardiometabolic health and slowed aging-related change in physiological-system integrity ^14,16,17^. Decades of follow-up will be required to determine if the CALERIE intervention affected disease onset or lifespan. In order to conduct an immediate human test of the geroscience hypothesis, we assayed DNA methylation from blood samples collected during the trial to quantify participants’ biological aging.

Biological aging is the progressive loss of system integrity that occurs with aging, driving disease, disability and mortality ^1,18^. There is no gold standard measure of biological aging; several methods have been proposed based on different biological measurements and analytic strategies ^19–21^. Among these, DNA methylation-based estimates of the state and pace of biological aging represent the current state of the art ^22^. Our analysis tested if CALERIE™ intervention slowed biological aging in the CR intervention group as compared to the Ad-libitum control group. We considered 3 groups of DNA methylation measures of biological aging: two first-generation DNA methylation clocks developed from analysis of chronological age by Horvath and by Hannum et al. ^23,24^; two second-generation DNA methylation clocks developed from analysis of mortality risk by Levine et al. (PhenoAge) and Lu et al. (GrimAge) ^25,26^; and two DNA methylation measures of the Pace of Aging developed from analysis of decline in system integrity, DunedinPoAm and DunedinPACE ^27,28^.

## METHODS

### Study design and participants

CALERIE Phase-2, was a multi-center, randomized controlled trial conducted at three clinical centers in the USA ^14^. It aimed to evaluate the time-course effects of 25% CR (i.e. intake 25% below the individual’s baseline level) over a 2-year period in healthy, adults (men aged 21–50 y, premenopausal women aged 21–47 y) with BMI in the normal weight or slightly overweight range (BMI 22.0-27.9 kg/m^2^). The study protocol (NCT00427193) was approved by Institutional Review Boards at three clinical centers (Washington University School of Medicine, St Louis, MO, USA; Pennington Biomedical Research Center, Baton Rouge, LA, USA; Tufts University, Boston, MA, USA) and the coordinating center at Duke University (Durham, NC, USA). All study participants provided written informed consent. Non-genomic data were obtained from the CALERIE Biorepository (https://calerie.duke.edu/apply-samples-and-data-analysis).

### Randomization and masking

After baseline testing, participants were randomly assigned at a ratio of 2:1 to a CR behavioral intervention or to an ad libitum control group (AL). Randomization was stratified by site, sex, and BMI. A permuted block randomization technique was used.

### Procedures

Study procedures were published previously ^14,16,29^ and are described here in brief. Participants in the CR group were prescribed a 25% restriction in calorie intake based on energy requirements estimated from two doubly-labelled water measurement periods at baseline. Participants were provided three meals per day for 27 days to familiarize themselves with portion sizes for a 25% reduced calorie intake; meals included eating plans modified to suit various cultural preferences. Participants also received instruction on the essentials of CR. Finally, participants were provided with intensive group and individual behavioral counselling sessions once a week with 24 group and individual counselling sessions over the first 24 weeks of the intervention. Adherence to the CR intervention was estimated in real time by the degree to which individual weight change followed a predicted weight loss trajectory (15.5% weight loss at 1 year followed by weight loss maintenance). The precise level of CR achieved was quantified retrospectively by calculating energy intake during the CR intervention and comparing it to baseline energy intake. Energy intake during the 2-year trial was quantified from total daily energy expenditure (assessed during 2-week doubly labeled water periods every 6 months) and changes in body composition (i.e., fat mass and fat-free mass). Participants assigned to the AL group continued on their regular diets; they received no specific dietary intervention or counselling. They had quarterly contact with study investigators to complete the assessments.

### DNA-methylation data

DNA extracted from blood samples was obtained from the CALERIE Biorepository at the University of Vermont. DNA methylation (DNAm) data were generated by the Kobor Lab at the University of British Columbia and processed by the Genomic Analysis and Bioinformatics Shared Resource at Duke University. Illumina Infinium Methylation EPIC BeadChip arrays were used to assay genome-wide DNA methylation data from banked DNA samples extracted from blood collected at the baseline, 12- and 24-month follow-ups. The EPIC array quantifies DNAm levels at >850,000 CpG sites across all known genes, regions, and key regulatory regions. Briefly, 750ng of extracted DNA samples were bisulfite converted using the EZ DNA Methylation kit (Zymo Research, Irvine, CA), and 160ng of the converted DNA were used as input for the EPIC arrays (Illumina, San Diego, CA). EPIC arrays were processed according to the manufacturer’s instructions and scanned using the Illumina iScan platform. To the extent possible, baseline, 12-month, and 24-month samples from the same individual were processed in the same array batch and on the same Beadchip to minimize batch effects; CR treatment and AL control participants were included on all chips. Quality control and normalization analyses were performed using the methylumi (v. 2.32.0) ^30^ Bioconductor (v 2.46.0) ^31^ package for the R statistical programming environment (v 3.6.3). Probes were considered missing in a sample if they had detection p-values>0.05 and were excluded from the analysis if they were missing in >5% of sample. Normalization to eliminate systematic dye bias in 2-channel probes was carried out using the methylumi default method. Following quality control and normalization, DNA methylation data for 828,613 CpGs were available for n=595 samples (baseline n=214; 12-months n=193; 24-months n=188). Additional batch correction was performed by residualizing DNA methylation measurements for principal components estimated from array control-probe beta values ^32^. Cell count estimation was performed using the Houseman Equation via the Minfi and FlowSorted.Blood.EPIC R packages ^33,34^.

### DNA methylation clocks and Pace-of-Aging measures

DNA methylation (DNAm) clocks are algorithms that combine information from DNAm measurements across the genome to quantify variation in biological age ^35^.

The first-generation of DNAm clocks were developed from machine-learning analysis comparing samples from individuals of different chronological age. These clocks were highly accurate in predicting the chronological age of new samples and also showed some limited capacity for predicting differences in mortality risk, although effect-sizes tend to be small and inconsistent across studies ^23,24,36^. We analyzed the first-generation clocks proposed by Horvath (Horvath Clock) and Hannum et al. (Hannum Clock) ^23,24^.

The second-generation DNAm clocks were developed with the goal of improving quantification of biological aging by focusing on differences in mortality risk instead of on differences in chronological age ^25,26^. These clocks also include an intermediate step in which DNA methylation data are fitted to physiological parameters. The second-generation clocks are more predictive of morbidity and mortality as compared to the first-generation clocks ^37^ and are proposed to have improved potential for testing impacts of interventions to slow aging ^38^. We analyzed the second-generation clocks proposed by Levine et al. (PhenoAge Clock) and Lu et al. (GrimAge Clock) ^25,26^.

A third generation of DNAm measures of aging are referred to as Pace-of-Aging measures. In contrast to the first- and second-generation clocks, which were developed by comparing DNAm data between individuals of different chronological ages, the Pace-of-Aging measures were developed from analysis of individuals who were all the same chronological age. Whereas the clocks aim to quantify how much aging has occurred up to the time of measurement, Pace of Aging measures aim to quantity how fast the process of aging-related deterioration of system integrity is proceeding. Pace-of-aging measures were developed by modeling within-individual multi-system physiological change over time in the Dunedin Study birth cohort ^39,40^. The first-generation pace-of-aging measure was based on analysis of change across three time points, when Dunedin Study members were aged 26, 32, and 38 years, and was named DunedinPoAm denoting “(P)ace (o)f (A)ging from DNA (m)ethylation” ^27^. The second-generation pace-of-aging measure was based on analysis of change across four time points, when Dunedin Study members were aged 26, 32, 38, and 45 years, and was named DunedinPACE for “(P)ace of (A)ging (C)omputed from the (E)pigenome” ^28^.

Additional details about the DNA methylation clock and Pace of Aging measures are reported in **Supplemental Table 1**.

### Analysis

We computed change-scores for the six aging measures by comparing values at the 12-month and 24-month follow-up assessments to baseline values (i.e. 12-month change = 12-month value – baseline, 24-month change = 24-month value – baseline). We conducted analysis of these change scores to test the hypothesis that CR slows biological aging using two complementary approaches: (1) we conducted Intent-to-Treat (ITT) analysis that compared change scores between participants randomized to CR-intervention and AL-control group; (2) we conducted Effect-of-Treatment-on-the-Treated (TOT) analysis using instrumental-variables methods to estimate the effect of CR on change scores.

In ITT analysis, we tested the effect of randomization to CR versus AL on aging-measure change scores using repeated-measures ANCOVA implemented under mixed models, following the approach used in past CALERIE analysis ^16^. The model included terms for treatment condition (CR or AL), follow-up time, an interaction term modeling heterogeneity in the treatment effect between the 12- and 24-month follow-ups, the baseline level of the aging measure, and the following pre-treatment covariates: chronological age, sex, race/ethnicity (Black, White, Other), body-mass index stratum at randomization (normal weight [22.0-24.9 kg/m2] and overweight [25.0-27.9 kg/m2]), and study site. Models were fitted using the Stata software’s ‘mixed’ command.

In TOT analysis, we tested the effect of the CR intervention on aging-measure change-scores using instrumental variables (IV) regression implemented using a two-stage least squares approach ^41^. The first stage regression modeled CR-treatment dose as a function of randomization condition (CR vs AL) and pre-treatment characteristics (chronological age, sex, race/ethnicity, body-mass index, study site, and baseline value of the biological-aging measure). The model instruments were randomization condition and interactions of randomization condition with sex and pre-treatment values of body-mass index and the biological-aging measure. The second stage regression modeled aging-measure change-scores as a function of the CR-treatment dose estimated from the first-stage regression and pre-treatment covariates. Separate models were fitted for the 12- and 24-month follow-ups. IV regression models were fitted using the Stata 16.0 software’s ‘ivregress’ command.

In ITT and TOT analysis, effect-sizes were scaled in standardized units according the distribution of the aging measures at pre-treatment baseline. For the DNAm clocks, clock-ages were differenced from chronological ages and standard deviations for these age-differences values were used for scaling. For the Pace of Aging measures, standard deviations of the original values were used for scaling. Treatment effects denominated in these standardized units are interpreted as Cohen’s d.

We conducted test-retest reliability analysis of DNAm measures of aging to investigate potential differences across measures. ITT and TOT analyses are based on analysis of change-scores computed from comparisons of repeated measures at two time points. Outcome-measures test-retest reliability therefore has strong influence on treatment-effect magnitudes. (Low test-retest reliability will bias effect estimates toward the null.) To inform interpretation of results, we computed test-retest reliability for the aging measures based on comparison of DNAm data generated for this study with DNAm data from a pilot study of the CALERIE baseline DNAm samples ^27^ using intraclass correlation coefficients (ICCs) ^42^. Analysis included n=183 samples included in both datasets. Aging measures were residualized for chronological age prior to analysis. The Horvath and Hannum clocks showed relatively lower reliabilities (ICC range 0.63-0.66), the PhenoAge, GrimAge, and DunedinPoAm measures showed intermediate reliabilities (ICC range 0.76-0.77), and the DunedinPACE measure showed higher reliability (ICC=0.89). We repeated this analysis using a publicly available dataset, GSE55763 ^32^ which included matched datasets for n=36 samples generated from Illumina 450k arrays. In that analysis, reliabilities were ICC=0.82, 0.85, 0.71, and 0.96 for the Horvath, Hannum, PhenoAge, and GrimAge clocks, respectively, and 0.89 and 0.96 for DunedinPoAm and DunedinPACE Pace of Aging measures.

## RESULTS

### Baseline characteristics in the CALERIE Trial

Of 238 eligible individuals, CALERIE randomized N=220 participants to treatment (145 CR-intervention and 75 AL-control; **Figure 1**). Blood DNAm data were generated at baseline and at least one follow-up timepoint for n=197 participants (128 CR and 69 AL). Of this analysis sample, n=105 (82%) of CR participants and n=59 (86%) of AL participants had DNAm data available from all three time points (baseline, 12-months, and 24-months). Participants had mean age of 38 years (SD=7), 70% were women, and 77% were white; there were no differences in age, sex, race/ethnicity between AL and CR at baseline (**Table 1**).

**Figure 1.**
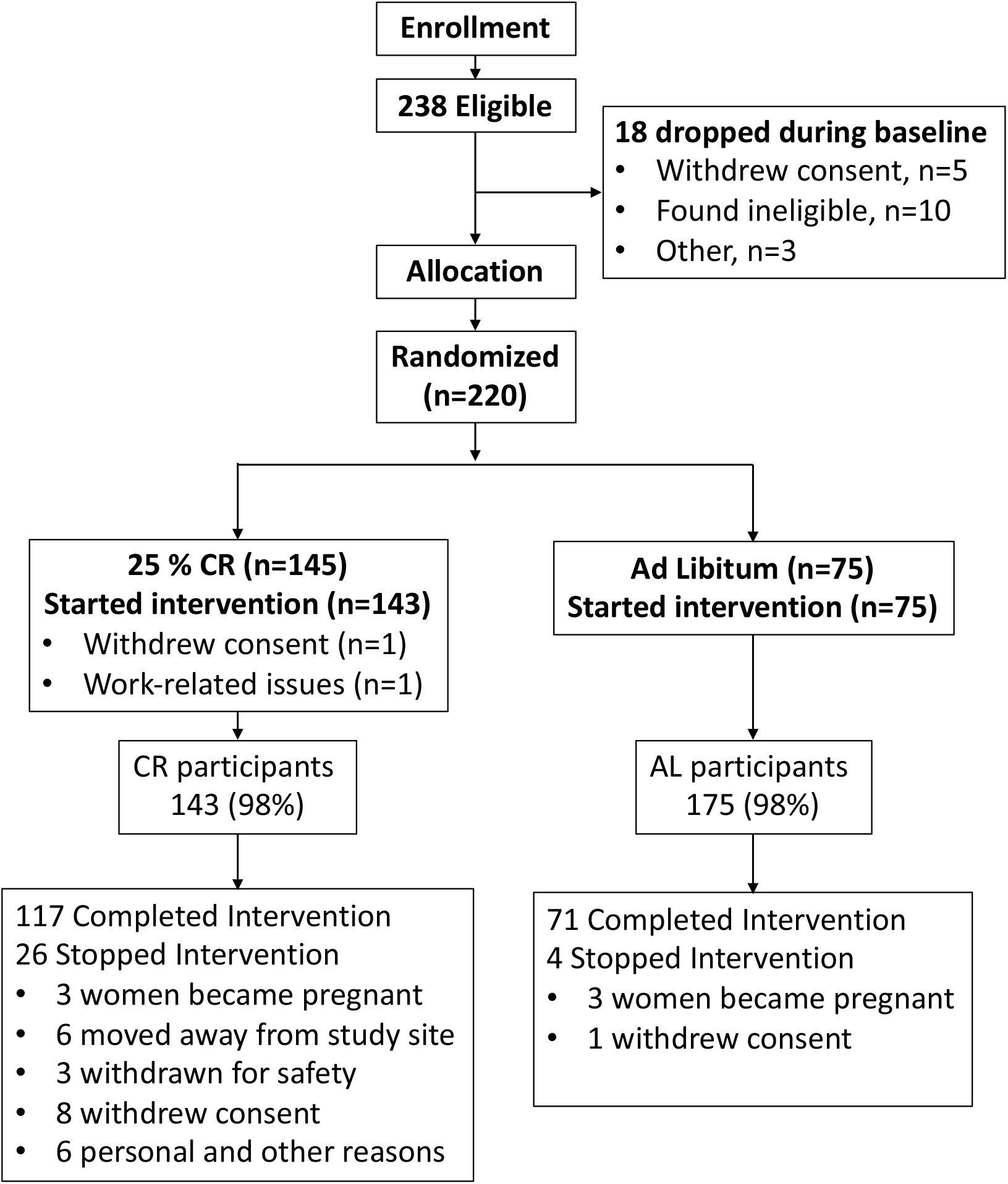
Consort Diagram for the CALERIE Trial. DNA methylation (DNAm) for analysis was assayed from blood samples collected at baseline, 12-months, and 24-months. Of the n=197 participants for whom DNAm data were available from baseline and at least one follow-up assessment, n=125 CR and 65 AL participants had DNAm data available at 12-months and n=117 CR and 65 AL participants had DNAm data available at 24 months.

**Table 1.**
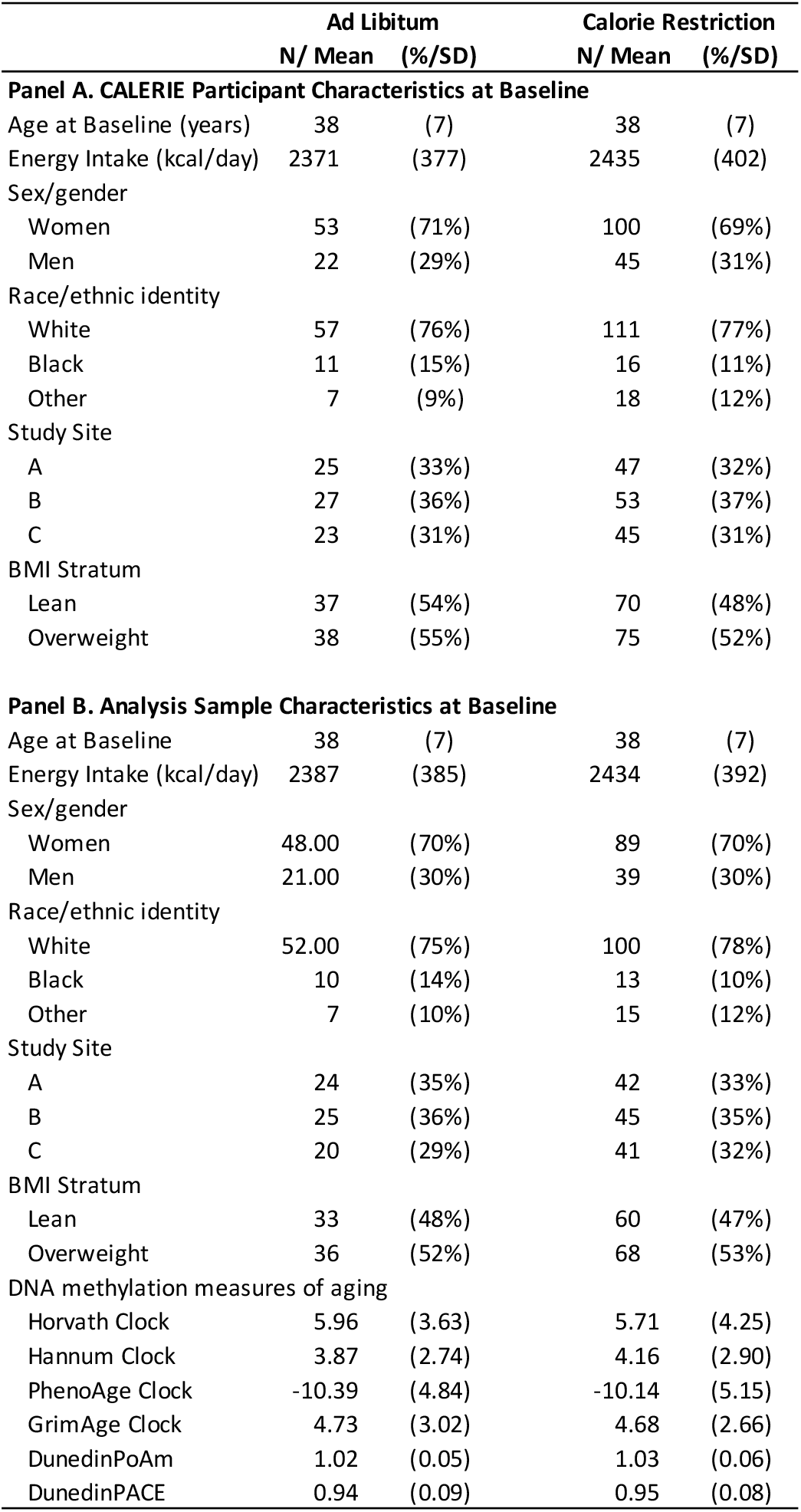
Characteristics of CALERIE trial participants at baseline. The table shows data for participants randomized to the ad libitum (AL) control group and the calorie restriction (CR) treatment group. CALERIE included a total of N=220 participants (AL n=75, of whom n=71 completed the study; CR n=145, of whom n=118 completed the study). The analysis sample was comprised of CALERIE participants for whom DNA methylation data were available at baseline and at least one follow-up assessment (“analysis sample”; N=197; AL n=69, CR n=128). Panel A reports characteristics of participants in the CALERIE Trial at baseline. Panel B reports characteristics of analysis sample participants at baseline.

Participants’ DNA-methylation-clock values were highly correlated with their chronological ages at baseline (r=0.83-0.92) and their Pace-of-Aging values were moderately correlated with their chronological ages at baseline (r=0.08 for DunedinPoAm; r=0.15 for DunedinPACE). Associations of aging measures with chronological age are graphed in **Supplemental Figure 1**. At baseline, participants’ clock-ages for the first-generation Horvath and Hannum clocks and the second-generation GrimAge clock were older than their chronological ages by 4-6 years; PhenoAge clock ages were younger than chronological ages by 10 years. CR and AL groups did not differ at baseline on the six aging measures although, for DunedinPoAm, the difference between CR and AL approached statistical significance at the alpha=0.05 level (d=0.24, p=0.120). Values of the six DNAm aging measures at baseline, and 12- and 24-month follow-ups in the CR and AL groups are reported in **Table 2**. Change-scores are reported in **Supplemental Table 2** and graphed in **Figure 2**.

**Table 2.** DNA-methylation clock and Pace of Aging values for CALERIE participants in the Ad Libitum (AL) and Caloric Restriction (CR) groups at baseline, 12-month, and 24-month assessments. Values for DNA methylation clocks are age-difference scores computed by subtracting participants’ chronological ages from their clock ages. DunedinPoAm and DunedinPACE Pace of Aging values are not adjusted for chronological age. 95% Confidence intervals are reported in brackets. Follow-up DNA methylation data for the n=69 AL and n=128 CR participants in the analysis sample were available for n=65 AL and n=125 CR participants at 12-months and n=68 AL and 117 CR participants at 24-months.

|  |  | **Ad Libitum (n=69)** |  | **Calorie Restriction (n=128)** |  |
| --- | --- | --- | --- | --- | --- |
|  |  | **Mean** | **95% CI** | **Mean** | **95% CI** |
| Horvath Clock | Baseline | 5.96 | [5.10 , 6.82] | 5.71 | [4.98 , 6.45] |
|  | 12mo | 5.41 | [4.46 , 6.37] | 5.45 | [4.74 , 6.15] |
|  | 24mo | 5.26 | [4.32 , 6.20] | 5.23 | [4.45 , 6.02] |
| Hannum Clock | Baseline | 3.87 | [3.22 , 4.51] | 4.16 | [3.66 , 4.67] |
|  | 12mo | 3.69 | [2.90 , 4.48] | 3.99 | [3.47 , 4.51] |
|  | 24mo | 3.85 | [3.14 , 4.56] | 4.02 | [3.47 , 4.57] |
| PhenoAge Clock | Baseline | -10.39 | [-11.53 , -9.24] | -10.14 | [-11.04 , -9.25] |
|  | 12mo | -10.28 | [-11.57 , -8.99] | -10.04 | [-10.98 , -9.10] |
|  | 24mo | -10.78 | [-11.90 , -9.65] | -10.13 | [-10.99 , -9.26] |
| GrimAge Clock | Baseline | 4.73 | [4.01 , 5.44] | 4.68 | [4.22 , 5.14] |
|  | 12mo | 4.59 | [3.87 , 5.30] | 4.13 | [3.68 , 4.59] |
|  | 24mo | 4.16 | [3.52 , 4.80] | 3.80 | [3.33 , 4.28] |
| Dunedin PoAm | Baseline | 1.019 | [1.008 , 1.031] | 1.032 | [1.022 , 1.042] |
|  | 12mo | 1.027 | [1.014 , 1.040] | 1.022 | [1.013 , 1.031] |
|  | 24mo | 1.017 | [1.006 , 1.029] | 1.024 | [1.014 , 1.034] |
| Dunedin PACE | Baseline | 0.940 | [0.919 , 0.962] | 0.946 | [0.932 , 0.960] |
|  | 12mo | 0.958 | [0.938 , 0.978] | 0.930 | [0.916 , 0.944] |
|  | 24mo | 0.953 | [0.934 , 0.971] | 0.931 | [0.915 , 0.946] |

**Figure 2.**
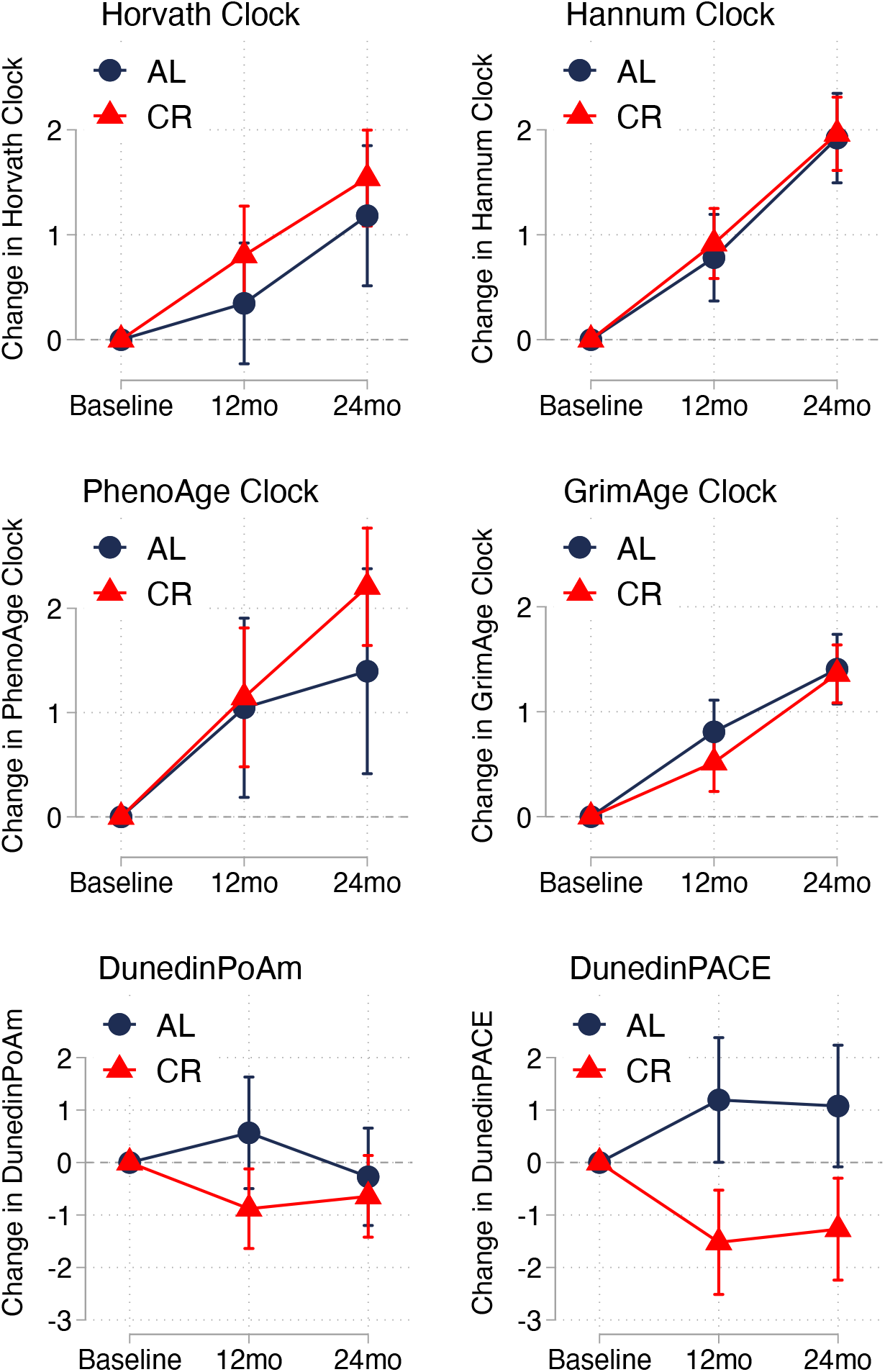
Change from baseline to 12- and 24-month follow-up in DNA methylation measures of aging in the ad libitum (AL) and caloric restriction (CR) groups in the CALERIE Trial. The figure graphs mean values and 95% confidence intervals of change-scores for the six DNA methylation measures of aging. Values for the AL control group are graphed in blue. Values for the CR treatment group are graphed in red. For the Horvath, Hannum, PhenoAge, and GrimAge DNA methylation clocks, change scores are denominated in “years” of DNA methylation age. For the clocks, expected change under the null hypothesis is 1 year at 12-months follow-up and 2 years at 24 months follow-up. For the DunedinPoAm and DunedinPACE measures, change scores are denominated in Pace of Aging units scaled to be interpretable as percent difference in the rate of aging relative to the reference norm of one year of biological decline per calendar year. For the Pace of Aging measures, expected change under the null hypothesis is zero.

### Intent-to-Treat (ITT) Analysis

For the first-generation Horvath and Hannum clocks and the second-generation PhenoAge clock, change over time did not differ between CR and AL groups and most associations were opposite the direction expected, i.e. change was faster in CR vs. AL (Cohen’s d effect-size range =-0.02 to 0.15, p>0.147). For the second-generation GrimAge clock, change in CR was slower as compared to AL, but the difference was not statistically different from zero at the alpha=0.05 level (12-month d=-0.12; 24-month d=-0.06; p>0.084).

For DunedinPoAm and DunedinPACE, intervention slowed Pace of Aging in the CR group as compared to the AL group, although differences were statistically significant only for DunedinPACE (for DunedinPoAm 12-month d=-0.19, 24-month d=0.01, p>0.101; for DunedinPACE, 12-month d=-0.29, 24-month d=0.24, p<0.002). ITT analysis effect-sizes are reported in **Table 3 Panel A** and graphed in **Figure 3 Panel A**.

**Table 3.** Effect-size estimates for intent-to-treat (ITT) and treatment-effect-on-the-treated (TOT) analysis testing the effect of CALERIE intervention on DNA methylation measures of aging. Panel A shows effect estimates for ITT analysis. Panel B shows effect estimates for TOT analysis. ITT analysis was conducted using mixed models to compare change-scores for DNA methylation measures of aging between CR and AL groups. Models included covariates for sex, race/ethnicity, study site, and pre-treatment baseline chronological age and body-mass index stratum. TOT analysis was conducted using instrumental variables regression to test associations of CR dose with aging-measure change-scores. Instruments included treatment group and interactions between treatment group and sex, body-mass index, and baseline levels of the aging measures. Model covariates were the same as in ITT analysis. TOT effect estimates are reported for a 20% dose of CR. This level of CR represents the 75^th^ percentile of the treatment group CR distribution at 12-months follow-up and the 87^th^ percentile of the treatment group CR distribution at 24-months follow-up. All effect-sizes are scaled in units equivalent to the standard deviation of the outcome measure at pre-intervention baseline. For DNA methylation clocks, standard deviations were computed from baseline age-difference values. For Pace of Aging measures, standard deviations were computed from untransformed baseline values.

|  |  | Treatment<br>Effect<br>(Cohen's d) | 95% CI | p-value |
| --- | --- | --- | --- | --- |
| **Panel A. ITT Effect Estimates (CR vs. AL)** |  |  |  |  |
| Horvath | 12mo | 0.10 | [-0.07, 0.27] | 0.255 |
| Clock | 24mo | 0.06 | [-0.12, 0.24] | 0.518 |
| Hannum | 12mo | 0.04 | [-0.14, 0.21] | 0.687 |
| Clock | 24mo | -0.02 | [-0.19, 0.15] | 0.804 |
| PhenoAge | 12mo | 0.03 | [-0.18, 0.23] | 0.784 |
| Clock | 24mo | 0.15 | [-0.05, 0.34] | 0.147 |
| GrimAge | 12mo | -0.12 | [-0.26, 0.02] | 0.084 |
| Clock | 24mo | -0.06 | [-0.20, 0.07] | 0.353 |
| Dunedin | 12mo | -0.19 | [-0.41, 0.04] | 0.101 |
| PoAm | 24mo | 0.01 | [-0.19, 0.21] | 0.937 |
| Dunedin | 12mo | -0.29 | [-0.45, -0.14] | 2.58E-04 |
| PACE | 24mo | -0.24 | [-0.39, -0.09] | 0.002 |
| **Panel B. TOT Effect Estimates (20% CR)** |  |  |  |  |
| Horvath | 12mo | 0.17 | [-0.05, 0.39] | 0.139 |
| Clock | 24mo | 0.07 | [-0.24, 0.37] | 0.672 |
| Hannum | 12mo | 0.08 | [-0.15, 0.31] | 0.498 |
| Clock | 24mo | -0.01 | [-0.33, 0.30] | 0.937 |
| PhenoAge | 12mo | -0.05 | [-0.32, 0.22] | 0.709 |
| Clock | 24mo | 0.17 | [-0.13, 0.48] | 0.262 |
| GrimAge | 12mo | -0.17 | [-0.37, 0.03] | 0.090 |
| Clock | 24mo | -0.09 | [-0.32, 0.13] | 0.419 |
| Dunedin | 12mo | -0.31 | [-0.61, 0.00] | 0.048 |
| PoAm | 24mo | -0.06 | [-0.39, 0.27] | 0.711 |
| Dunedin | 12mo | -0.41 | [-0.64, -0.18] | 4.73E-04 |
| PACE | 24mo | -0.35 | [-0.61, -0.08] | 0.011 |

**Figure 3.**
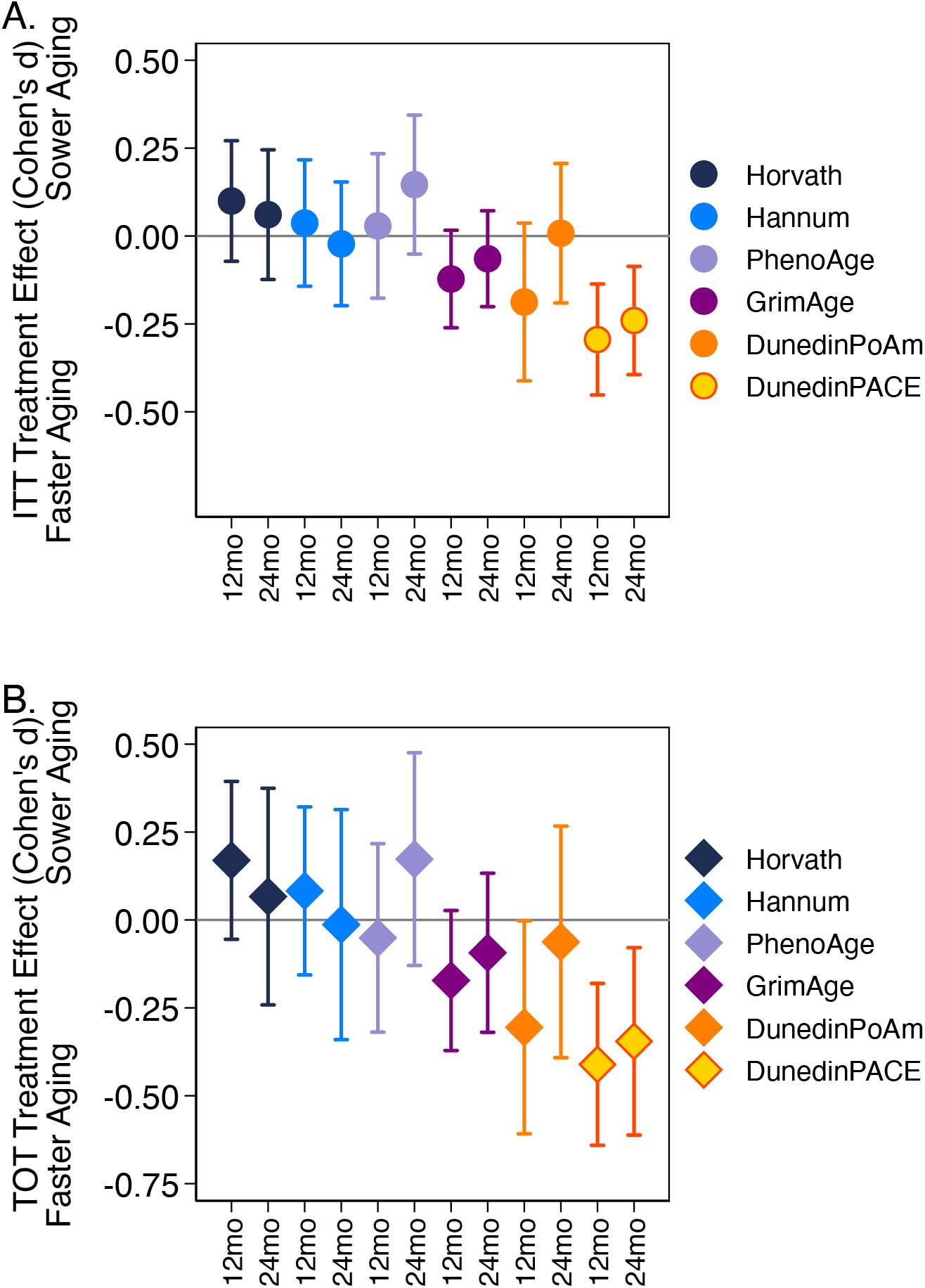
Effect-sizes for Intent-to-Treat (ITT) and Treatment Effect on the Treated (TOT) analysis of DNA methylation measures of aging in the CALERIE Trial. Panel A plots ITT treatment effects estimated from mixed models testing association of random assignment to the CR intervention group with change scores for the aging measures. ITT effect-sizes represent standardized differences in change scores between ad libitum control and calorie restriction treatment groups at the 12- and 24-month follow-up assessments. Panel B plots TOT treatment effects estimated from two-stage least-squares instrumental variables regression testing association of percent calorie restriction with change scores for the aging measures. TOT effect-sizes represent standardized effects of 20% calorie restriction on change scores of the aging measures at 12- and 24-month follow-up assessments. ITT and TOT effect-sizes are denominated in pre-intervention-baseline standard-deviation units of the outcome measures, interpretable as Cohen’s d. Error bars show 95% confidence intervals.

Treatment effect-size magnitudes were correlated with test-retest reliabilities for DNAm measures of aging (r>0.6). Measures with higher ICCs had larger and more consistent effect-sizes at 12- and 24-months **Supplementary Figure 2**.

We conducted additional analysis to evaluate dose-response in treatment effects. We repeated analysis using the same regression models used to estimate the ITT effects, with stratification of the treatment group by percent CR achieved (>/<10%). Results indicated larger effects for those achieving higher doses of CR. Analysis of CR treatment effects with the intervention group stratified by CR dose are reported in **Supplemental Table 3**.

### Effect-of-Treatment-on-the-Treated (TOT) Analysis

TOT analysis results were similar to results from ITT analysis and analysis in which the CR-treatment group was stratified by CR dose. For the first-generation Horvath and Hannum clocks and the PhenoAge clock, CR was not associated with delayed biological aging and most associations were opposite the direction expected, i.e. higher CR dose was associated with more-rapid increase in clock-age (Cohen’s d effect-size range =-0.05 to 0.17, p>0.139). For the second-generation GrimAge clock, higher CR dose was associated with slower increase in clock-age, but the effect was not statistically different from zero at the alpha=0.05 level (12-month d=-0.17; 24-month d=-0.09; p>0.090).

For the Pace of Aging measures, higher CR dose was associated with reduced Pace of Aging, although the 24-month effect for DunedinPoAm was not statistically different from zero at the alpha=0.05 level (for DunedinPoAm 12-month d=-0.31 p=0.048, 24-month d=-0.06 p=0.711; for DunedinPACE, 12-month d=-0.41, 24-month d=-0.35, p<0.011). TOT analysis effect-sizes are reported in **Table 3 Panel B** and graphed in **Figure 3 Panel B**.

### Sensitivity Analyses

We tested sensitivity of results to covariate adjustment for estimated cell counts by repeating analyses including estimated cell counts as model covariates. Results were similar to results from unadjusted analyses (**Supplementary Table 4**).

We tested sex differences in treatment effects. Means of aging measures and aging-measure change-scores stratified by sex are reported in **Supplemental Table 5**. We repeated ITT and TOT analysis with the addition of a product term testing interaction between the treatment variable and participant sex. Sex differences in treatment effects were not statistically different from zero in any of the models. Sex-stratified treatment effects and tests of sex differences in treatment effects are reported in **Supplemental Table 6**.

## DISCUSSION

In animal experiments, CR modifies molecular hallmarks of aging, reduces risk factors for many aging-related diseases, and extends healthy lifespan, suggesting that this intervention slows biological aging ^5,10–13^. Here we report DNA methylation (DNAm) evidence that CR may also slow biological aging in humans. We analyzed DNAm measures of biological age and Pace of Aging at pre-treatment baseline and at 12- and 24-month follow-ups in a randomized controlled trial of CR in healthy, non-obese humans, CALERIE™. We found that CR slowed Pace of Aging, especially for the DunedinPACE measure, and modestly delayed increases in biological age according to the GrimAge DNA methylation clock. In contrast, the clocks proposed by Horvath, Hannum et al., and Levine et al. (PhenoAge) did not show this effect and sometimes suggested an effect in the opposite direction.

Our findings contrast with reports that aging rates may not be modifiable ^43^ and advance proof-of-concept that an intervention proven to extend healthy lifespan in animals can slow the pace of biological aging in humans. They contribute gold-standard randomized-controlled-trial evidence to early reports from small and un-controlled intervention studies that DNAm measures of aging can be modified in humans ^38,44,45^. And they extend to the molecular level of analysis in our prior finding in CALERIE™ that CR slows the rate of increase in blood-chemistry biological age ^17^.

The significance of DNAm evidence of slowed Pace of Aging/ delayed biological aging in CALERIE is remains to be established. Epigenetic alterations, including changes to DNAm, are among the hallmarks of aging ^2,3,46^. The DNAm measures of aging we studied are predictive of a range of healthspan metrics in young, midlife, and older adults, including morbidity and mortality ^25,27,28,47,48^. However, the extent to which changes to DNAm measures of aging in CALERIE reflect CR-induced changes to hallmarks of aging is not certain. And additional follow-up of trial participants is required to determine whether CR-induced slowing of biological aging in CALERIE will translate into disease prevention and increased healthy lifespan.

CR effects varied substantially across the DNAm measures of aging we analyzed. Overall, Pace of Aging measures were more sensitive to intervention as compared to clock measures. Within these categories, DunedinPACE Pace of Aging was more sensitive than DunedinPoAm and GrimAge was more sensitive as compared to the other clocks. This pattern of findings mirrors differences in how the measures were developed and in their measurement reliability.

The DNAm measures of aging we studied were developed using a common approach but are based on distinct models of aging. The common approach was elastic-net regression machine-learning analysis ^49^ applied to Illumina-array whole-genome DNA methylation data. However, the targets of machine learning analysis, and therefore the underlying models aging, were different. The first-generation Horvath and Hannum clocks used chronological age differences between individuals as their target. The second-generation PhenoAge and GrimAge clocks used between-individual differences in mortality risk as their target. Both targets represent the accumulated effects aging across the entire lifespan. In contrast, the Pace of Aging measures used as their target differences between individuals who were all the same chronological age in the rate of decline in system integrity (measured over 12 years of follow-up for DunedinPoAm and 20 years of follow-up for DunedinPACE). This target represents an estimate of how fast aging is proceeding in the years leading up to DNAm measurement. One possible explanation for why Pace of Aging measures were more sensitive to intervention as compared to the clocks is that the clocks, because they summarize aging related changes accumulated over the entire lifespan, may be less sensitive to interventions that slow aging, but do not substantially reverse it.

Measurement reliability is a key consideration in studies of change in response to treatment. The aging measures we studied varied in their test-retest reliability, with DunedinPACE and GrimAge showing higher reliabilities as compared to the other measures. Test-retest reliability ICC statistics for the aging measures were correlated with their treatment-effect magnitudes; the more-reliable measures had larger treatment effects. This result suggests that new methods proposed to improve reliability of DNAm measures of aging may reveal still-larger treatment effects in CALERIE and have potential to increase statistical power of other trials utilizing these measures as endpoints ^50,51^.

We acknowledge limitations. There is no gold standard measure of biological aging ^52^. We analyzed several proposed DNA methylation measures of aging, which are recognized as the best available measurements of this construct ^53^. Nevertheless, these measures are acknowledged to be incomplete summaries of biological changes that occur with aging and to have technical limitations, including imperfect test-retest reliability ^50,51,54,55^. Treatment-effect estimates may therefore represent a lower-bound of the true impact of CALERIE intervention on aging. The measures of aging we studied summarize biological aging in general and do not isolate system-specific aging processes ^56^. However, CR has diverse effects on aging across multiple biological systems ^57,58^. Our general measures of aging thus provide a reasonable test of cross-system impacts. On average, trial participants did not achieve the prescribed dose of 25% CR and some control-group participants reduced their caloric intake. Despite this imperfect adherence, treatment-group participants experienced substantial and sustained weight loss and related changes in body and tissue composition, broad improvement in cardiometabolic health, and a slowing of aging-related physiological changes ^16,17,59,60^. Our TOT analysis indicated that participants who achieved higher doses of CR experienced more pronounced reductions in Pace of Aging and slowed rate of increase in the GrimAge clock. The CALERIE Trial sample does not represent the general population. Treatment effects may not generalize beyond the population of healthy volunteers recruited to participate. CALERIE follow-up is, so far, limited to the end of the intervention period. Whether treatment and any slowing in biological aging that resulted from it translated to long-term clinical benefit is currently unknown.

Within the context of these limitations, our findings have implications for future research. Aging biology research has identified multiple therapies with potential to improve healthy lifespan in humans. A barrier to advancing translation of these therapies through human trials is that intervention studies run for months or years, but human aging takes decades to cause disease ^61–63^. New measurements that summarize biological changes occurring with aging have potential to overcome this challenge; measurements to quantify biological aging that both predict future disease, disability, and mortality and can detect changes in aging processes over short time scales have potential function as surrogates for intervention effects on healthy lifespan ^22,53^. The several methods proposed to quantify biological aging analyzed in this study are predictive of aging-related health decline and mortality. However, until this study, none had been tested in a randomized controlled trial (RCT) of a geroscience-based intervention ^22^. Our findings highlight two measures with potential utility in future trials, the GrimAge clock and the DunedinPACE Pace of Aging. These two measures have the highest test-retest reliability and show the strongest associations with healthspan endpoints in validation studies ^25,28,51^. CALERIE trial evidence contributes further validation that these measures are appropriate for testing geroscience interventions in humans.

Findings also illustrate challenges facing future trials. CALERIE was a 24-month, intensive behavioral intervention to deliver a therapy proven to slow aging in animal models. Even so, treatment effect-sizes were modest. While even modest slowing the Pace of Aging can have profound effects on population health ^64–66^, future trials, especially those considering less-intensive or shorter-term interventions, such as intermittent fasting ^67^, should plan for larger samples and focus on the most reliable measurements. Models forecasting returns from interventions to delay aging ^64^ may be most informative when focused on relatively modest effects of intervention. Ultimately, establishing DNA methylation measures of aging as surrogate endpoints for geroscience will require trials that include long-term follow-up to establish intervention impacts on primary healthy-aging endpoints, including incidence of chronic disease and mortality ^68–70^. The evidence reported from CALERIE suggests that some DNA methylation measures of aging may be helpful in identifying those interventions most promising for such long-term follow-up.

## Data Availability

All non-genomic data are available from the CALERIE biorepository (calerie.duke.edu). Genomic data are available from corresponding author until March 2024, at which point they will be available from the CALERIE biorepository.

http://calerie.duke.edu

## Acknowledgements

This research was supported by R01AG061378. Additional support for VBK was provided by R01AG054840. We thank the CALERIE Research Network R33AG070455 for their assistance in this project and The Dunedin Study R01AG032282 for facilitating early access to the DunedinPACE DNA methylation algorithm.

DLC and DWB are listed as inventors on a Duke University and University of Otago invention that was licensed to a commercial entity.

